# Anxiety, depression, attitudes, and internet addiction during the initial phase of the 2019 coronavirus disease (COVID-19) epidemic: A cross-sectional study in México

**DOI:** 10.1101/2020.05.10.20095844

**Authors:** Bryan Adrián Priego-Parra, Arturo Triana-Romero, Samanta Mayanin Pinto-Gálvez, Cristina Durán Ramos, Omar Salas-Nolasco, Marisol Manriquez Reyes, Antonio Ramos-de-la-Medina, Jose María Remes-Troche

## Abstract

**Objectives:** To describe the prevalence and distribution of anxiety and depression among Mexican population, and to examine its association with internet addiction during the COVID-19 outbreak.

**Design:** A web-based cross-sectional study.

**Setting:** General population in México.

**Participants:** 561 subjects were recruited (71% female, mean age 30.7 ± 10.6 years).

**Interventions:** An online survey to assess personal attitudes and perceptions towards COVID-19, sleep-disorders related, the Mexican version of the Hospital Anxiety and Depression Scale (HADS) and the Internet Addiction Test (IAT) was applied.

**Primary and secondary outcome measures:** Prevalence of anxiety, depression, internet addiction and sleep disorders and associated factors. Also, prevalence for anxiety and depression were compared to an historic control group.

**Results:** During the initial phase of the COVID-19 pandemic the prevalence for anxiety and depression was 50% (95% CI, 45.6% to 54.1%) and 27.6%, (95% CI 23.8% to 31.4%), respectively. We found a 51% (33% to 50%) increase in anxiety and up to 86% increase in depression during the initial weeks of the lock-down compared to the control group. According to the IAT questionnaire, 62.7% (95% CI 58.6% to 68.8%) of our population had some degree of internet addiction. Odds ratio for development of anxiety symptoms was 2.02 (95% CI1.56-2.1, p=0.0001) and for depression was 2.15 (95% CI 1.59-2.9, p=0.0001). In the multivariate analysis, younger age (p=0.006), sleep problems (p=0.000), and internet addiction (p=0.000) were associated with anxiety and depression.

**Conclusions:** Our study provides valuable information on the psychological impact that the COVID-19 pandemic has had on the Mexican population. As in other parts of the globe, in Mexico, fear of SARS-CoV-2 infection has had devastating consequences on mental health, such as anxiety, depression and sleeping disturbances. Internet abuse and the consequent overexposure to rapidly spreading misinformation (infodemia) are associated to anxiety and depression.

Strengths and limitations of this study

- Our study have addressed the immediate psychological effect of the pandemic in the general population in a Latin American country, specifically in Mexico, a nation with high population density.
- Using the IAT (a specific tool to assess internet dependency), we found internet addiction was highly prevalent and correlated to anxiety and depression.
- We used the snowball sampling strategy; thus, our population is biased and may not reflect the actual pattern of general population.
- We decided to compare anxiety and depression with an historic cohort, and although this control group is not exactly matched to our studied population, the prevalence of anxiety and mood disorders are like those reported previously in Mexico.
- Other limitations include response bias due to fewer older subjects participating, the fact that sleep problems were not rigorously evaluated with a specific tool, and some states in our country were not represented in this work.

## Introduction

In early December 2019, the first cases of an unknown type of pneumonia in humans were reported in Wuhan City, Hubei Province, China.^1^ The World Health Organization (WHO) working along with the Chinese authorities identified the etiological agent as a new type of coronavirus which was first named novel coronavirus (nCoV-2019) and finally severe acute respiratory syndrome coronavirus (SARS-CoV-2).^2^ Over a period of a few weeks, the infection spread across the globe at rapid pace and on January 30^th^, the WHO declared a public health emergency of international concern.^3^ On February 11st, 2020 the novel virus disease was named Coronavirus Disease – 19 or “COVID-19”. On March 11^th^, COVID-19 was declared a pandemic.^4^

COVID-19 has been since its origins a highly contagious disease, the virus spread rapidly across the planet, and by early May 2020 it had infected more than 3.3 million people in 187 countries..^5^ Due to its high infection rate, lethality and lack of previous immunity, this novel infection has been perceived as a major threat to the life and health of the global human population.

The first case of COVID-19 in Latin America was reported in Brazil on February 26th, 2020, and the first death on March 7th in Argentina.^6^ In Mexico, the first case was reported on February 25^th^ and the first death on March 18^th^.^8^ Mexico is the 13th-largest country in the world in size and the 10^th^ most populous with 128,649,565 inhabitants.^7^ The Mexican government declared a national health emergency on March 30th and implemented restrictions in the public, private, and social sectors which include: voluntary quarantine (lock-down), school closings, social distancing, and limitation of non-essential activities.^9^

As in other parts of the globe, life in Mexico has dramatically changed at many different levels and fields: family, science, medicine, economics, religion, culture, human behavior, and technology.^10^ It’s known that both massive tragedies and global disease outbreaks (like COVID-19) have a negative impact on mental health. Previous studies have established a clear link between pandemic and symptoms of stress, depression, anxiety, post-traumatic stress, and suicidal tendencies.^10-12^ According to the behavioral immune system (BIS) theory, personal reactions to the COVID-19 pandemic, include the development of self-protection mechanisms, such as fear, negative emotions, and negative cognitive evaluation.^13^

In addition to the fear and anxiety caused by the virus itself, several other factors may negatively impact the mental health of people under lock-down. ^14,15^ The internet has become an essential and inseparable part of the modern lifestyle.^16^ However, there is a “problematic behavior of human interactions with information and communication technologies” that has led to the development of concerning long-term issues.^17^ The term “internet addiction,” which is defined as “a psychological dependence on the internet, regardless of the type of activity once logged on,” describes this problematic behavior.^18^ Several studies have found that internet addiction increases the risk of depression, anxiety and stress.^16-19^

Due to the sudden onset of COVID-19, few studies have explored the effects of SARS-COV-2 on mental health.^14,20,21^ To our knowledge there are no studies in Latin American countries that have explored how this pandemic may increase internet addiction, and its relationship to triggering anxiety and depression in the general population. We believe that a comprehensive management of the pandemic is essential, not only focusing on the physical aspects and infected patients, but also on mental health, which can be directly reflected in ideas, emotions, and cognition.

Our study aims to describe the prevalence and distribution of anxiety and depression among Mexican population, and to examine its association with internet addiction during the COVID-19 outbreak, using a rapid internet-based assessment.

## Methodology

### Study design, setting and sampling

A cross-sectional study was conducted online from March 23^th^ to April 21th, 2020, when increased numbers of COVID-19 cases were identified, along with an increased potential for person-to-person transmission in Mexico (phase 2). Mexican citizens aged >18 years old were invited to participate in an online survey using the Google Forms software. The link to the questionnaire was sent through email and social media. The participants were encouraged to roll out the survey to as many people as possible (snowball sampling),thus the link was forwarded to people apart from the first point of contact and so on. Upon receiving and clicking the link, participants got auto directed to information on the study and informed consent.

### Data collection and study procedure

The online survey included 30 items, divided into five categories: a) demographic data (age, gender, state of origin, comorbidities, previous diagnosis of anxiety and depression), b) personal attitudes and perceptions towards COVID-19 (beliefs, level of trust in prevention recommendations, symptoms, and when to search medical care), c) sleep-disorders related questions, d) the Mexican version of the Hospital Anxiety and Depression Scale (HADS) ^22,23^ and finally, e) the Internet Addiction Test (IAT).^24^ Subjects with a previous diagnosis (in the past 3 months) of anxiety and depression were excluded.

HADS has been widely used because its good performance in identifying caseness and assessing symptom severity for anxiety disorders and depression in somatic, psychiatric, and primary care patients, and in the general population. HADS has been validated in Spanish for the Mexican population.^22,23^ HADS consists of two subscales: one measuring anxiety (HAD-A), with seven items, and another measuring depression (HAD-D), also with seven items, which score separately. Each item is answered by the patient on a 4-point (0-3) scale, so the possible scores range from 0 to 21 for each of the two subscales, taking 2-5 minutes to complete. The original authors post a score of ≥11 as indicating the presence (“caseness”) of a mood disorder, and a score of 8-10 being just suggestive of the presence of the respective state.^22^ As proposed by others, we considered a score of ≥8 as positive for either anxiety or depression symptoms.^25^ The sensitivity and specificity of HADS-A and HADS-D with a threshold of 8+ is often found to be in the range of 0.70 to 0.90 with areas under the curve (AUC) of 0.84 −0.96.^22^

The IAT Spanish version is a 20-item scale that measures the presence and severity of internet dependency among adults,^26^ each item consists of a Likert-scale with a range of 0 to 5. According to the score at the end of the test, patients can be classified into four categories: 0 to 19 can be considered as absence of addiction, 20 to 39 indicates a low level of addiction and average online user, 40 to 69 a moderate level of addiction, and a score ≥70 a severe level of internet addiction.^24,26^

### Ethical considerations

Data were protected according to the General Data Protection Regulation. The text message bearing the link to the Google Form which was shared with the participants contained information on the title and aim of the study, the eligibility of participants, information on the benefits and harm, and the average duration required to fill a questionnaire which was 3 min. This information was also found on the first page of the Google Form and participants were given the option to either consent to participate (which will give them access to the questionnaire) or decline to participate (which will automatically submit a blank form). This study was approved by the Internal Institutional Review Board of the Medical Biological Research Institute (IIIMB-UV#2020-01-005).

### Statistical analysis

Descriptive statistics have been used in this study to analyze the findings. Mean and standard deviation and proportions have been used to estimate the results of the study. The χ2 /trend tests were used to determine the prevalence of depression, anxiety, and combination of depression and anxiety by categorical variables including social internet addiction and sleep problems. Multivariate logistic analysis regression was used to explain the association between the prevalence of depression, anxiety, and internet addiction. We estimated the adjusted ORs and their 95% confidence intervals (CIs) of independent variables for frailty. The IBM SPSS version 24 (IBM, Chicago, Illinois, US) was used to carry out all analyses.

As a control group, to compare anxiety and depression symptoms, we used data from 458 subjects (62% female, mean age 29.8 ± 8.6 years) to whom the same questionnaires for anxiety and depression were applied between September 2019 and January 2020. These subjects come from a study of prevalence of anxiety and depression in open population that is ongoing in our country as part of a screening program for metabolic syndrome and non-alcoholic fatty liver disease.

## Results

### Sociodemosraphic characteristics

We received responses from 593 participants, 22 were excluded because reported a previous anxiety or depression diagnosis. Thus, 561 subjects were analyzed, 400 were female (71%), and the mean age was 30.7 ± 10.6 years. There were respondents from 25 out of the 32 states that constitute México. All respondents were from urban areas.

### Attitudes, beliefs and perceptions about COVID-19 pandemic

Most of the participants (99.6%) were aware of the pandemia and consider SARS-COV2 as a real threaten to their health. Eighty two percent (n=459) perceive themselves at risk to get infected and develop COVID-19 during the following months. Most (98%) of the participants thought social distancing was essential to stop the virus from spreading. Most participants (97%) acknowledged that washing hands frequently could stop the spread of infection.

Figure 1 shows what would be the behavior of subjects in case of acquiring COVID-19. Although 82% of respondents would attend ER if presenting alarm signs, 14% will attend as soon if they had the first symptom that for them was suspicious for COVID-19. Most of the respondents will accept hospitalization and all required measures (95%, n=532) in case of a complicated clinical course; 5% (n=29) then, consider that there are no useful treatments for severe cases and fatality is inevitably highly. One hundred and eighty-one (33%) consider that the fear and worries related to COVID-19 has interfered with the quality of their sleep (Figure 2).

**Figure 1.**
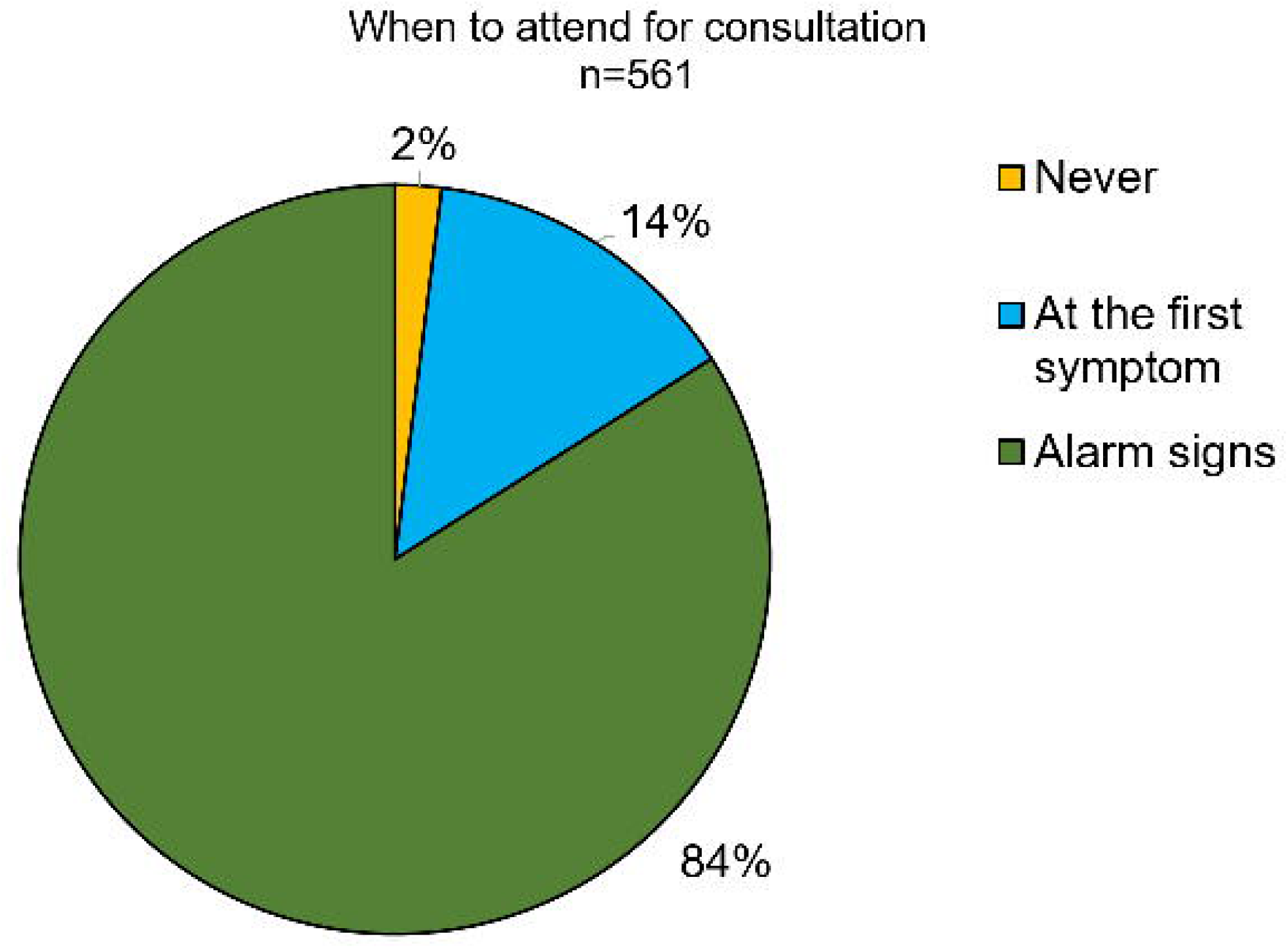
- Attitude of Mexican population about when to attend for consultation in case of acquiring CVODI-19.

**Figure 2.**
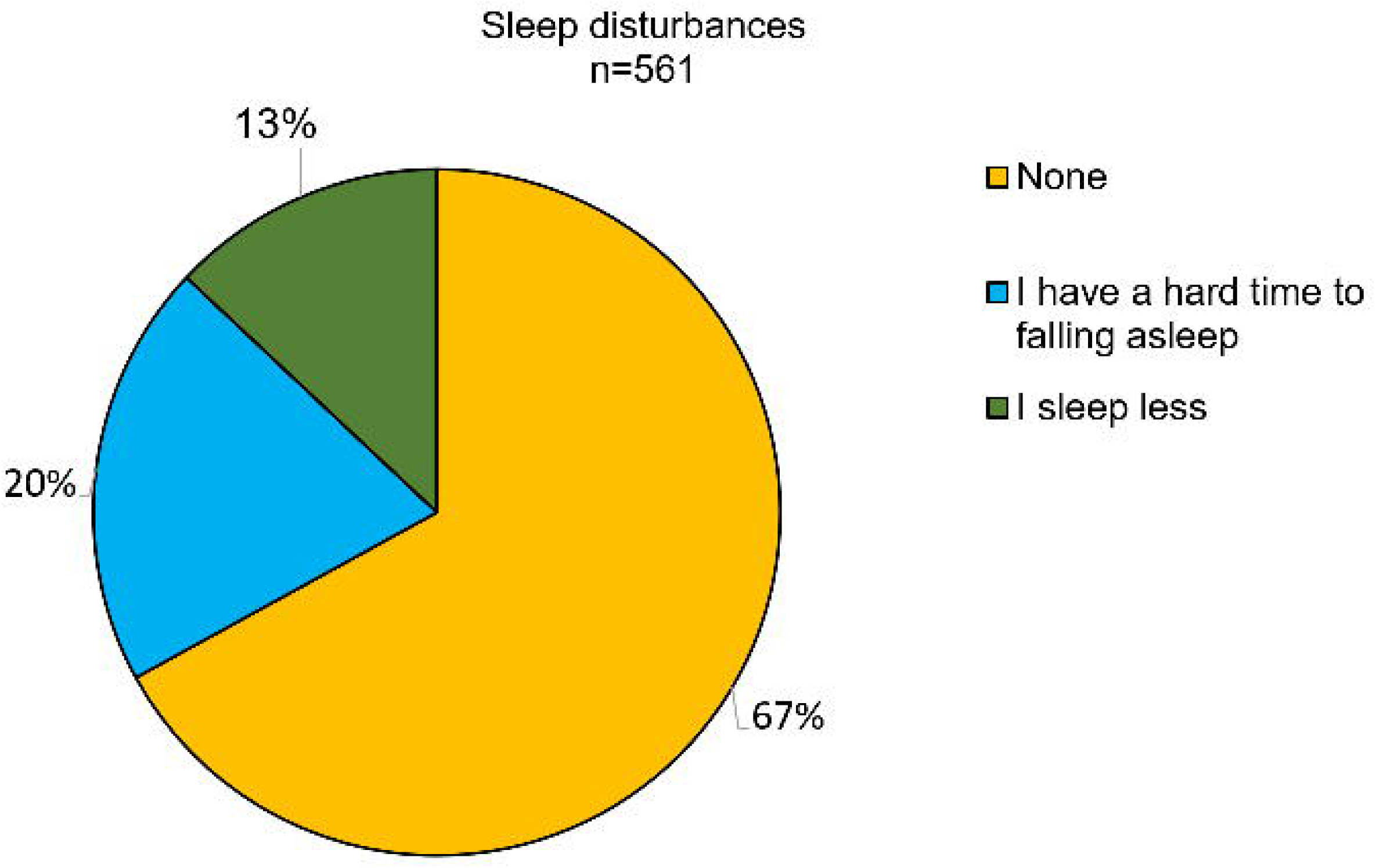
- Prevalence of sleep disturbance (%) among surveyed subjects.

### Anxiety and Depression according to HAD questionnaire

Two hundred and eighty subjects (50%; 95% CI, 45.6% to 54.1%) have developed anxiety symptoms during the COVID-19 breakout (Figure 3). According to the different cut-off levels proposed for the HAD questionnaire there were 115 subjects (21%; 95% CI, 17.0 % to 23.9%) with possible anxiety disorder and 165 (29.5%; 95% CI, 25.5% to 36.2%) with the presence of anxiety disorder. Regarding depression, 155 subjects (27.6%, 23.8% CI to 31.4%) have developed depression symptoms during the phase 2 of the COVID-19 pandemic. Ninety-six (17.1%, 95% CI 13.9% to 20.3%) have a possible depression disorder and 59 (10.5%, 95% CI 7.8% to 10.1%) have presence of a depression disorder. One hundred and forty-two subjects (25.3%, 95% CI 21.6% to 28.9%) have overlapping anxiety and depression symptoms.

**Figure 3.**
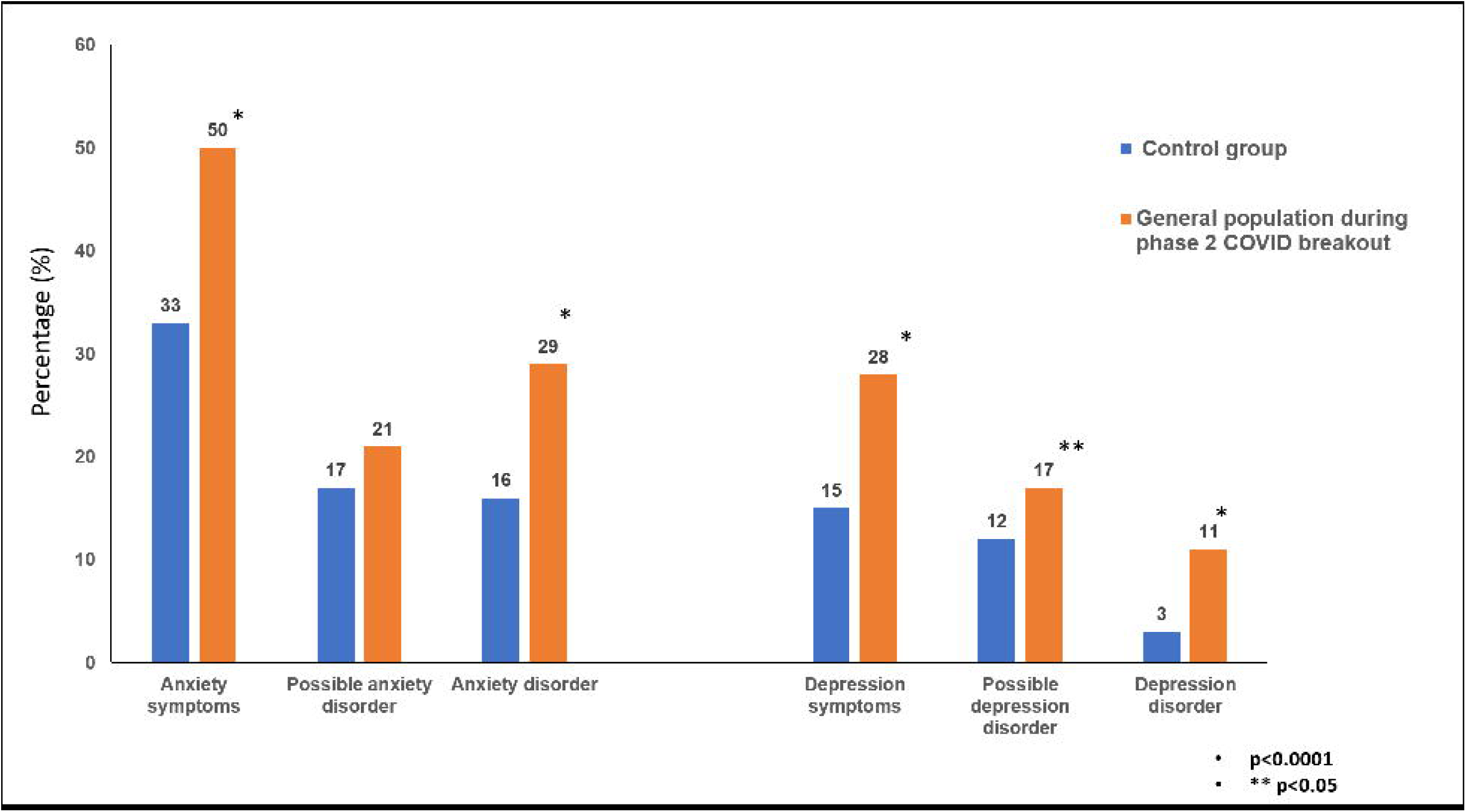
- Percentage of subjects with anxiety and depression during the COVID-19 pandemic compared to a historic control group. Anxiety or depression symptoms were considered if subjects had a score >8 in the HAD. Those subjects whose scores were between 8-10 were classified as “suspect”, meanwhile those with a score > 11 were considered as a caseness.

### Internet addiction

According to the IAT questionnaire, 352 (62.7%, 95% CI, 58.6% to 68.8%) of our surveyed population had some degree of internet addiction: 293 (52.2%, 95% CI 48.0% to 64.8%) low, 57 (10.2%, 95% CI 7.5% to 12.7%) moderate and 2 (0.4%, 95% CI 0.04% to 1.2%) severe addiction. Subjects with internet addiction were younger and had higher scores for anxiety and depression (Table 1). Also, prevalence for anxiety, depression and sleep disturbance were higher in subjects with higher levels of internet addiction (Table 1).

**Table 1.**
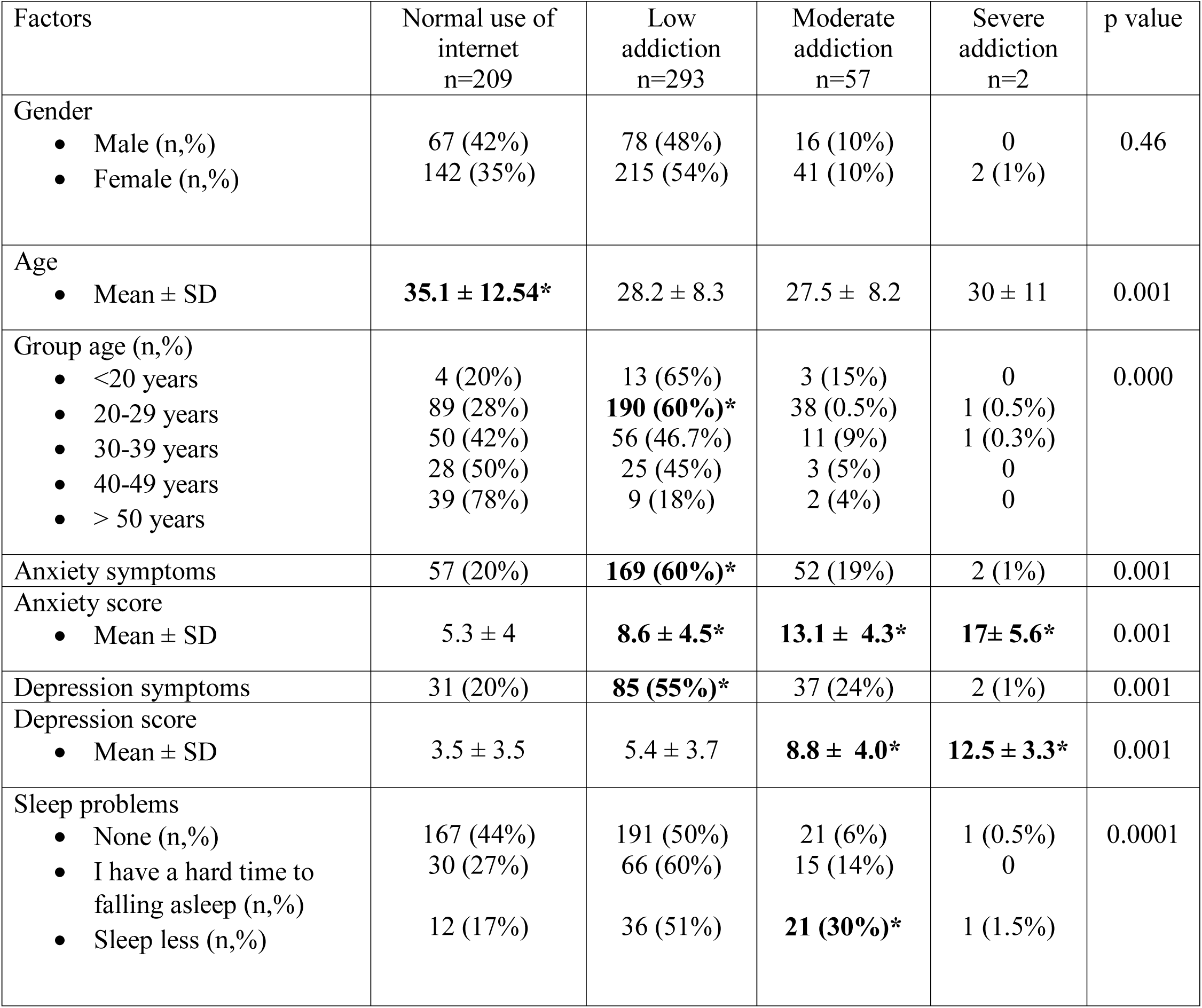
Demographic characteristics and factors associated to internet addiction.

### Prevalence of anxiety, depression and internet addiction compared to the control group

As is shown in Figure 3, compared to the control group, the prevalence for anxiety, depression and internet addiction was significantly higher in the participants that were evaluated during the initial phase of the COVID-9 pandemic. Odds ratio for development of anxiety symptoms was 2.02 (95% CI 1.56-2.1, p=0.0001), for possible anxiety disorder was 1.2 (95% CI 0.9-1.7, p=0.19) and for anxiety disorder was 2.1 (95% CI 1.6-2.9, p=0.0001) during the lock-down compared to control group. In the case of depression, OR for depression symptoms was 2.15 (95% CI 1.59-2.9, p=0.0001), for possible depression disorder was 1.41 (95% CI 1.01-2.06, p=0.04) and for depression disorder was 4.03 (95% CI 2.3-8.1, p=0.0001).

### Factors associated to anxiety, depression, and correlations

Univariate analysis found that female gender (80% vs 63%, p=0.0001), younger age (28.7 ± 8.1 vs. 32.6 ± 12.1, p=0.001), sleep disturbances (51.8% vs 12.8%,p=.0001) and a higher proportion of internet addiction (79.6% vs 45.9%, p=0.001) were associated among anxiety subjects (Table 1). Same factors were associated to depression (Table 2). Subjects with overlapping anxiety and depression (n=142, 25%) reported the highest scores for anxiety (13.3± 3.5) and depression s (10.6 ± 2.4)

**Table 2.** Factors associated to anxiety symptoms

| Factors | Anxiety<br>n=280 | No anxiety<br>n=281 | p value | OR<br>(95% CI) |
| --- | --- | --- | --- | --- |
| Gender<br>Male (n,%)<br>Female (n,%) | 57 (20%)<br>**223 (80%)\*** | 104 (37%)<br>177 (63%) | 0.0001 | 2.29 (1.5-3.3) |
| Age<br>Mean ± SD | **28.7 ± 8.1\*** | 32.6 ± 12.3 | 0.001 | ----- |
| Group age (n,%)<br><20 years<br>20-29 years<br>30-39 years<br>40-49 years<br>> 50 years | 9 (45%)<br>176 (55%)<br>63 (54%)<br>24 (44%)<br>8 (16%) | 11 (55%)<br>143 (45%)<br>54 (46%)<br>31 (56%)<br>**42 (50%)\*** | 0.000 | 1.0000<br>1.2 (0.7-2.0)<br>1.2 (0.7-1.9)<br>0.9 (0.5-1.7)<br>0.3 (0.16-0.7) |
| Sleep problems<br>None (n,%)<br>I have a hard time to falling asleep (n,%)<br>Sleep less (n,%) | 135 (48.2%)<br>**84 (30%)\***<br>**61 (21.8%)\*** | 245 (87.2%)<br>27 (9.6%)<br>9 (3.2%) | 0.0001 | 0.5 (0.4-0.6)<br>3.2 (2.0-4.7)<br>6.7 (3.3-13.6) |
| Internet addiction<br>None<br>Low<br>Moderate<br>Severe | 57 (20.4%)<br>**169 (60.4%)\***<br>**52 (18.6%)\***<br>2 (0.7%) | 152 (54.1%)<br>124 (44.1%)<br>5 (1.8%)<br>0 | 0.0001 | 0.3 (0.2-0.5)<br>1.3 (1-1.7)<br>10.4 (4.1-26)<br>----- |

**Table 3.** Factors associated to depression symptoms

| Factors | Depression<br>n=155 | No depression<br>n=406 | p= | OR |
| --- | --- | --- | --- | --- |
| Gender<br>Male (n,%)<br>Female (n,%) | 25 (16.1%)<br>**130 (83.9%)\*** | 136 (33.5%)<br>270 (66.5%) | 0.0001 | 2.6 (1.6-4.2) |
| Age<br>Mean ± SD | 28.7 ± 8.1 | 31.4 ± 11.3 | 0.002 | ----- |
| Group age (n,%)<br><20 years<br>20-29 years<br>30-39 years<br>40-49 years<br>> 50 years | 5 (25%)<br>94 (29%)<br>39 (33%)<br>12 (22%)<br>5 (10%) | 15 (75%)<br>225 (71%)<br>78 (67%)<br>43 (78%)<br>45 (90%) | 0.024 | 1.0000<br>1.17 (0.5-2.5)<br>1.3 (0.5-2.9)<br>0.8 (0.3-2.1)<br>0.4 (0.12-1.23) |
| Sleep problems<br>None (n,%)<br>I have a hard time to falling asleep (n,%)<br>Sleep less (n,%) | 66 (42.6%)<br>48 (31%)<br>41 (26.5%) | **314 (77.3%)\***<br>63 (15.5%)<br>29 (7.1%) | 0.0001 | 0.2 (0.1-0.2)<br>0.7 (0.5-1.1)<br>1.4 (0.8-2.2)<br>----- |
| Internet addiction<br>None<br>Low<br>Moderate<br>Severe | 31 (20%)<br>85 (54.8%)<br>37 (23.9%)<br>2 (1.3%) | 178 (43.8%)<br>208 (51.2%)<br>**20 (4.9%)\***<br>0 | 0.0001 | 0.1 (0.1-0.2)<br>0.4 (0.3-0.5)<br>1.8 (1.1-3.1)<br>----- |

Anxiety and depression scores have a good positive correlation (r=0.746). Anxiety scores had a moderate positive correlation with IAT scores (r=0.548), but weak negative correlation with age (r=-0.21). Similar, depression scores had a moderate positive correlation with IAT scores (r=0.51), but weak negative correlation with age (r=-0.19).

In the multivariate lineal regression model, younger age (Beta coefficient= −0.097, p=0.006), sleep problems (Beta coefficient= 0.221, p=0.000), internet addiction (Beta coefficient= 0.196, p=0.000) and concomitant depression (Beta coefficient=0.356, p=0.000) were associated with anxiety. For depression, the factors associated in the multivariate model were sleep problems (Beta coefficient= 0.135, p=0.001), internet addiction (Beta coefficient=0.104, p=0.011) and concomitant anxiety (Beta coefficient= 0.44, p=0.000).

## Discussion

Mental health and psychological disorders continue to be a problem that have a great impact in the quality of life of people and a high economic cost for the individual, society, and health systems.^27^ Public health emergencies (infectious and non-infectious) such as H1N1 outbreak,^28^ earthquakes,^29^ terrorist attacks,^30^ SARS,^31^ Ebola,^32^ and recently, COVID-19,^33,34^ can cause, trigger or worsen mental health problems,.

Fear and anxiety related to epidemics and pandemics negatively influence the behavior of people in the community. The new coronavirus outbreak has provided an important field for research in mental health in the last months. Despite the short time since the COVID-19 |pandemic started, there are already some studies (most of them carried out in China) that demonstrate the behavior and immediate psychological effect of the pandemic in the general population. So far in Latin America, the psychological impact of this new threat to our physical and mental health is unknown.

From the time the Mexican government identified an expanded potential for person-to-person transmission of SARS-CoV-2 (phase 2) and an accelerated growth in COVID-19 cases, anxiety and depression significantly increased in Mexico. To assess this situation we used HADS because it’s validated in Spanish and performs well in screening the caseness and different dimensions of anxiety and depression among non-psychiatric hospital patients as well as in the general population.^25^ We found a 51% (33% to 50%) increase in anxiety symptoms (HAD-A >8) and up to 86% (15% to 28%) increase in depression symptoms (HAD-D >8) during the initial weeks of the lock-down compared to the control group. If we consider a cut-off value of >11, the increase in anxiety disorders was 81% (15% to 29%) and 266% in depression (3% to 11%).

The last national mental health survey conducted in Mexico, showed that the overall prevalence for anxiety disorders was 14.3%,^35^ and that 4.5% of the population may suffer from some mood disorder, including depression. These percentages are similar to those found in our control group (15% and 3% respectively), thus, we believe that the increase we are reporting in anxiety and depression is real and reflects how the COVID-19 pandemic triggers anxiety and depression in our population. This is an important and remarkable finding considering that the knowledge and attitude towards COVID-19 in our population was high in terms of awareness of the disease, and measures to avoid infection such as social distancing and washing hands.

Our results are consistent with those reported in other populations during the outbreak. Wang et al.,^21^ in a study that included 1210 respondents from 194 cities in China, found that 53.8% of respondents rated the psychological impact of the outbreak as moderate or severe; 16.5% reported moderate to severe depressive symptoms, and 28.8% reported moderate to severe anxiety symptoms. Gao et al ^14^ in a study conducted with 4872 participants from 31 provinces and autonomous regions in China, found that prevalence of depression, anxiety and combination of depression and anxiety was 48.3% (95%CI: 46.9%-49.7%), 22.6% (95%CI: 21.4%-23.8%) and 19.4% (95%CI:18.3%-20.6%) during the COVID-19 outbreak in Wuhan, China. A third Chinese study also showed presence of anxiety symptoms in 35% and depressive symptoms in 20.1% of their population.^36^

Several factors have been associated to anxiety and depression during the COVID-19 pandemic in other countries such as: female gender, student status, poor self-rated health status and frequent social media exposition. In our study, using the IAT (a specific tool to assess internet dependency), we found that low to mild levels of internet addiction were highly prevalent (62%) and correlated to anxiety and depression.^14,21,36^ To our knowledge, this is a novel finding not previously reported. Gao et al,^14^ found that during the pandemic, 82.0% of the Chinese population was frequently expose to social media. Internet abuse is very likely to be associated with overexposure to rapidly spreading misinformation (infodemia) and unfounded fears due to the fact that people constantly express their negative feelings, worries, and anxiety on social media, which may have a contagious effect. Overuse of internet to obtain information about COVID-19 may exacerbate stress responses, increase levels of anxiety and depression, amplify concern, and impair functioning. This can lead to additional media consumption and further distress, creating a cycle that can be difficult to break.^37^

According to our results internet abuse was also associated to sleep problems and younger age. Roy et al,^20^ found that, in Indian population, sleep difficulties, paranoia about acquiring COVID-19 infection and distress related to social media use were reported in 12.5 %, 37.8 %, and 36.4 % participants, respectively. The negative impact of internet abuse on both sleep duration and sleep quality has been previously described in the literature.^38^ For example, Kim et al ^39^ found that Korean subjects with internet addiction were 1.7 times more likely to experience poor sleep quality in comparison to those with normal internet use. Also, subjects who spent more time in the web have more (2.5 times higher) risk of developing depressive symptoms. Our results are also consistent with previous reports that the younger generations trend to be heavier internet users and are more exposed to social media exposition than older people.^40^ The reasons are multiple but mainly involve technical skills and availability of internet resources.

An important factor associated to anxiety and depression in our study was younger age. This is a surprising finding considering that young people are considered a low risk group for complications and mortality associated with SARS-CoV-2. It has even been described that the possibility of being an asymptomatic carrier of the virus is greater in young people than in older subjects.^41^ One explanation is that younger generations, who are at home as consequence of the COVID-19 outbreak, may be experiencing stressful situations to which they have never been exposed before, such as changes in their routines, uncertainty, concern for their health and insecurities about the impact and duration of the disease. The lack of social interaction has caused an increase in mental health problems as well. Panic, stress, anxiety, depression, boredom, anger, suicidal ideation, and sleep disturbances have also been reported as a consequence of social isolation.^42,43^

In addition, staying home for long periods of time can lead to receiving less sunlight, which could decrease serotonin levels. This has been associated with emotional disorders like anxiety but mainly depression.^44^ As in other studies, we found that women were more likely to develop anxiety and depressive symptoms during the COVID-19 outbreak compared to men.^36^ This finding corresponds with extensive epidemiological studies which have found that women are, in general, at higher risk of developing depression.^45^ “Metaworry”, the persistent worry about one’s own thoughts and cognitive processes, is more frequent in women than in men. Thus, it is possible that fear to COVID-19 triggers a negative metacognitive process in which one worries about one’s own worrying which could potentially cause harm to oneself.^46^

It is important to acknowledge that our study has several limitations. As in other studies performed during this period, we adopted the snowball sampling strategy, thus our population is biased and may not reflect the actual pattern of general population. In an ideal scenario we should conduct a prospective evaluation of the same group of participants a period after the pandemic. We acknowledge that additional longitudinal studies, such as cohort studies or nested case-control studies are essential in the future. However, we decided to compare anxiety and depression with an historic cohort, and although this control group is not exactly matched to our studied population, the prevalence of anxiety and mood disorders are like those reported previously in Mexico. Other limitations include response bias due to fewer older subjects participating, the fact that sleep problems were not rigorously evaluated with a specific tool, and some states in our country were not represented in this work.

In conclusion, our study provides valuable information on the psychological impact that the COVID-19 pandemic has had on the Mexican population. COVID-19 has brought major changes in our lives and interpersonal relationships, creating uncertainty, fear, confusion, panic. In Mexicans, it has specifically had devastating consequences on mental health, such as anxiety, depression and sleeping disturbances. Federal agencies and some academic institutions (such as Universidad Veracruzana) have implemented several support groups intended for the open population to have prompt access to resources to deal with the effects of the critical events we are experiencing. At these difficult times for all human kind, it’s necessary to rapidly to increase health education and identify mental health disruptors to establish, early effective measures to lessen the impact of COVID-19 outbreak on our mental health.

## Data Availability

The dataset analyzed to generate the findings for this
study are available from the corresponding author on reasonable request.

## Patient consent for publication

Not required.

## Data availability statement

The dataset analyzed to generate the findings for this study are available from the corresponding author on reasonable request.

## Author Statement

Conception of study: JMRT. Data collection: BAGP, ATR, SMPN, CRD. Data curation and analysis: BAGP, JMRT. Interpretation of results: JMRT, MMR, OSN, ARDLM First draft of manuscript: JMRT, BAGP. Manuscript revisions: JMRT, MMR, OSN, ARDLM. Critical revision of the manuscript: JMRT, ARDLM. All authors read and approved the final version of the manuscript.

## Acknowledgements

The authors extend their sincere gratitude to Ms. Maria Medellin for her help for having translated and revised the grammar of this manuscript.

## ORCID iD

Jose María Remes-Troche https://orcid.org/0000-0001-8478-9659

